# Bovhyaluronidase azoximer for long-term pulmonary sequelae of COVID-19: a randomized, double-blind, placebo-controlled trial

**DOI:** 10.1101/2024.09.19.24313792

**Authors:** Sergey N. Avdeev, Galina L. Ignatova, Oxana M. Drapkina, Veronica B. Popova, Ekaterina V. Melnikova, Tatiana I. Chudinovskikh, Olga V. Ryabova, Natalia V. Egorova, Tamara V. Rubanik, Yury G. Shvarts, Svetlana A. Polyakova, Vitalina Dzutseva, Anna V. Antonova, Dmitry A. Zubkov, Mikhail S. Khmelevskii, Nadezhda F. Khomyakova, Mikhail A. Tsyferov, Tim C. Hardman, Anton A. Tikhonov

**Affiliations:** Department of Pulmonology, I.M. Sechenov First Moscow State Medical University (Sechenov University), Moscow, Russia; Department of Therapy, South Ural State Medical University, Chelyabinsk, Russia; National Medical Research Center for Therapy and Preventive Medicine, Moscow, Russia; Pavlov First Saint Petersburg State Medical University, St. Petersburg, Russia; Medical Center for Diagnostics and Prevention LLC, Yaroslavl, Russia; Department of Hospital Therapy, Department of Hospital Therapy, Kirov State Medical University, Kirov, Russia; City Clinical Hospital №2, Izhevsk, Russia; Energy of Health LLC, Saint Petersburg, Russia; Pulmonary department, City Consultative and Diagnostic Centre №1, St. Petersburg, Russia; Saratov State Medical University, Saratov, Russia; Revma-Med Medical Center LLC, Kemerovo, Russia; NPO Petrovax Pharm LLC, Moscow, Russia; Niche Science & Technology, Richmond, UK

**Author notes:** **Corresponding Author:** Sergey N. Avdeev, Department of Pulmonology, I.M. Sechenov First Moscow State Medical University (Sechenov University), 8 Trubetskaya Street, Postal Code 119991, Moscow, Russia. **Take-Home Message:** Bovhyaluronidase azoximer improved exercise tolerance in patients with post-COVID-19 pulmonary sequelae. Benefit to pulmonary function was observed in high-risk groups.

## Abstract

**Background:** Hyaluronan is an emerging target for COVID-19 and lung fibrosis. In an open-label study the hyaluronidase bovhyaluronidase azoximer (BA) was associated with improved pulmonary function and exercise tolerance in patients with pulmonary sequelae of COVID-19. In this randomized, double-blind, placebo-controlled trial we evaluated the effect of BA on patients up to 12 months after COVID-19, characterized by reduced pulmonary function, dyspnea, and decreased oxygen saturation.

**Methods:** Patients (n=392) were randomized 1:1 to receive BA (3000U) or placebo every 5 days for 71 days. Percent of predicted forced vital capacity (ppFVC), respiratory symptoms, and exercise tolerance indicators were assessed at baseline and on days 71 and 180. The primary endpoint was a change from baseline in ppFVC by Day 71.

**Results:** On Day 71, BA was associated with a significant reduction in the proportion of patients with exertional desaturation (OR=0.35, p=0.0051) and dyspnea (OR=0.62, p=0.043). There were no significant intergroup differences in the ppFVC growth rate. Analysis of sub-populations revealed that by Day 180, BA was associated with increased ppFVC in patients with cardiovascular comorbidities (diff=3.31%, p=0.042) and those with earlier SARS-CoV-2 infection (diff=4.17%, p=0.021). BA was generally safe and well-tolerated.

**Conclusion:** In patients with long-term pulmonary sequelae of COVID-19, BA was associated with increased exercise tolerance. There was evidence of shorter pulmonary function recovery time following BA in patients with cardiovascular comorbidities and those with earlier COVID-19 disease.

## Introduction

Long COVID or post-acute COVID-19 syndrome, includes a range of persistent and often debilitating symptoms affecting multiple organ systems. According to the World Health Organization, approximately 10–20% of COVID-19 patients develop long COVID. Estimates suggest that 65 million people worldwide suffer from long COVID, although actual prevalence may be higher [1]. Common and persistent sequelae include respiratory and cardiovascular symptoms, as well as general symptoms such as fatigue and exercise intolerance [2,3]. Long COVID manifestations are often associated with radiological lung abnormalities, indicating fibrotic-like changes, and restrictive pulmonary disease, collectively referred to as COVID-19 pulmonary sequelae [4,5].

SARS-CoV-2 infection triggers lung tissue fibrotic remodeling, vascular injury, and chronic inflammation, leading to the development of pulmonary sequelae [4,6,7]. Apart from the fibrotic response, these mechanisms also contribute to non-pulmonary symptoms of long COVID [1]. Hyaluronic acid (HA) is a molecular target involved in these processes. Hyaluronic acid and its metabolites mediate inflammatory response and fibrotic tissue remodeling and are implicated in several chronic inflammatory and fibrotic conditions. Multiple animal studies have demonstrated that targeting HA through hyaluronidases can ameliorate fibrosis and halt pathological inflammatory response [8,9,10].

Hyaluronic acid is abundant in the lungs of patients with COVID-19 and may contribute to acute respiratory distress syndrome (ARDS) development through fluid accumulation, inflammatory response and impaired gas exchange [11,12]. High serum HA levels have been observed in patients with more severe forms of COVID-19 infection and in cases where patients demonstrated associated organ damage. Clinical trial data suggests that inhibition of HA synthesis by 4-methylumbelliferone leads to a reduction in pulmonary lesions in patients with SARS-CoV-2 infection [13]. Furthermore, HA fragments generated in the lungs of COVID-19 patients directly induce vascular injury [14].

The role of HA in inflammatory and fibrotic conditions, and acute COVID-19 highlights the need for further investigation into its potential involvement in the pulmonary sequelae of COVID-19. Bovhyaluronidase azoximer (BA) is a PH20 hyaluronidase conjugated to a chemical polymer to prolong its half-life. A recent open-label study in patients with pulmonary sequelae subsequent to COVID-19 infection indicated that BA was associated with improvements in pulmonary function, exercise tolerance, and respiratory symptoms [15]. We conducted a randomized, double-blind, placebo-controlled clinical trial to further evaluate the possible effects of bovhyaluronidase azoximer (BA) on COVID-19 pulmonary sequelae.

## Methods

### Study design and oversight

We conducted a randomized, double blind, placebo-controlled, parallel-group clinical trial to assess the effect of BA on long-term pulmonary sequelae of COVID-19. The study was conducted at 37 sites. Patients were assigned randomly (1:1 ratio) to receive either BA (3000U) or a matching placebo intramuscularly every 5 days for 71 days (15 injections). We aimed to enroll 196 patients per trial group. After treatment completion patient entered a 109-day follow-up. Randomization was performed using an interactive web-response system.

Patients were screened within a 14-day period prior to randomization (Day 1). Study assessments were conducted at screening and on days 1, 31 (±1), 71 (±1), and 180 (±3). The following procedures were performed: spirometry, lung diffusion test, 6-minute walking test (6-MWT), pulse oximetry, and dyspnea evaluation (Modified Medical Research Council (mMRC) and Borg scales). Quality of life was assessed using the European Quality of Life 5-Dimension 5-Level Questionnaire (Euro-QoL-5D-5L) and cough severity was assessed using a visual analog scale (VAS). Chest computer tomography (CT) was performed at screening; CT scans performed during the screening period but outside of the study procedures were also permitted. Baseline values correspond to screening or Day 1 measurements, whichever was later.

The protocol was reviewed and approved by Russian Ministry of Health, and by an independent ethics committee at each participating center. The study was conducted in accordance with the Declaration of Helsinki and good clinical practice. All patients provided written informed consent before entering the study. The study was registered with ClinicalTrials.gov, number NCT06383819.

### Patient population

We enrolled patients aged 18–80 years with a history of COVID-19 infection within 1 to 12 months of the screening, reporting pulmonary post-COVID-19 sequelae manifested by residual lung abnormalities, restrictive pulmonary disease, dyspnea, and decreased oxygen saturation at rest or after exercise. Eligible participants had a predicted forced vital capacity (FVC) <80% with a forced expiratory volume in one second (FEV1) to FVC ratio >70%, and residual lung abnormalities affecting more than 10% of lung tissue, including ground glass opacities, consolidations, and other abnormalities indicative of previous COVID-19 as assessed by a specialist. Chest CT scans were assessed locally. Further eligibility criteria included dyspnea of >1 point on the mMRC scale; oxygen saturation (SpO2) at rest below 95%; or desaturation defined as a ≥4% decrease in SpO2 after 6-MWT.

The exclusion criteria were chronic respiratory diseases, other conditions that can lead to restrictive pulmonary disease, dyspnea of etiology not linked to post-COVID-19 condition, serious cardiac disorders within 6 months before screening, acute infections, and preexisting liver or kidney disease.

### End points

Change from baseline in the percent of predicted FVC (ppFVC) by Day 71 was the primary endpoint. Secondary endpoints related to FVC were the change from the baseline in ppFVC by Day 180 and the proportions of patients achieved at least 10% ppFVC increase by days 71 and 180.

Other secondary endpoints included changes from baseline in SpO2 at rest, cough severity (visual analog scale) and quality of life (Euro-QoL-5D-5L); the proportion of patients with SpO2 at rest ≤93%; the proportion of patients exhibiting desaturation after the 6-MWT; the proportion of patients with exertional dyspnea (defined by a ≥2 point increase on the Borg scale after 6-MWT); the proportion of patients achieving a ≥50 meter increase in distance walked during the 6-MWT compared to the distance at baseline; and the proportion of patients with a decrease in resting dyspnea from baseline by ≥1 point on the mMRC scale. All secondary endpoint measures were assessed on days 71 and 180.

Adverse event severity and causality were assessed by investigators and reviewed by the study medical monitor.

### Statistical analysis

Intergroup comparisons for the primary endpoint were performed using an ANCOVA model, with the treatment group as a factor and baseline value as a continuous covariate. Statistical significance for the primary endpoint was determined at a two-sided 5% level. For secondary endpoints representing the proportions of patients, intergroup comparisons were performed using generalized linear models, treating these endpoints as dichotomous variables. Continuous secondary endpoints were analyzed using the same approach as the primary endpoint. Missing data was imputed based on the average of non-missing data from three similar patients within the same treatment group. Patients were considered similar if they had no missing values at previous visits and exhibited the smallest sum of squared deviations from the corresponding values at the previous visit.

## Results

First patient randomization occurred on April 28, 2022, and the last patient completed the trial on September 4, 2023. Patients (n=392) were randomized and received at least one dose of BA or placebo, 382 completed the trial. All randomized patients were included in the safety population. The Full Analysis Set (FAS) included 376 patients (186 in the BA group and 190 in the placebo group: Fig. 1). The primary reason for exclusion from the FAS was non-compliance with the inclusion criteria of restrictive pulmonary disease (FVC <80%, FEV1/FVC >70%).

**Figure 1.**
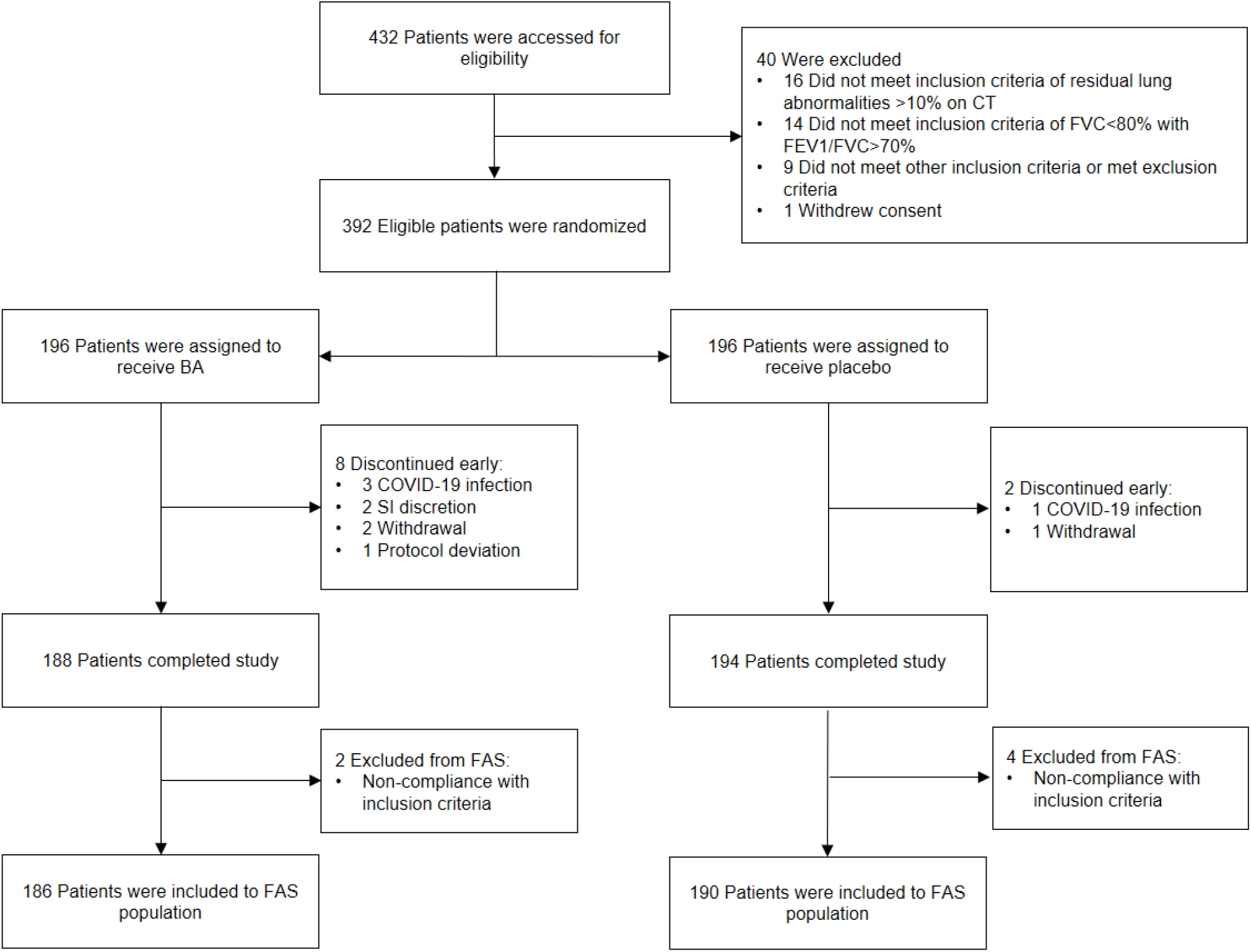
CONSORT flow diagram FAS: full analysis set; SI: site investigator.

Baseline demographic and clinical characteristics were similar in BA and placebo groups (Table 1), although there were slightly fewer females in the BA group. Patients in the BA group tended to have a lower percent lung damage, less exertional dyspnea, and greater distances in the 6-MWT.

**Table 1.** Patient demographics and baseline characteristics*.

|  | **BA (N=186)** | **Placebo(N=190)** |
| --- | --- | --- |
| Male sex - number (%) | 78 (42) | 69 (36) |
| Age | 55.7±11.7 | 58.5±11.2 |
| Body mass index | 29.5±5.0 | 29.7±5.2 |
| Time from Covid-19 start and randomization date | 220±105 | 217±112 |
| Covid-19 hospitalization status |  |  |
| - Hospitalized – number (%) | 127 (68) | 135 (71) |
| - Not hospitalized – number (%) | 59 (32) | 55 (29) |
| Cardiovascular comorbidities – number (%) | 140 (75) | 141 (74) |
| Percent of predicted FVC | 69.6±7.3 | 69.0±7.8 |
| Percent of lung damage | 18.5±11.5 | 21.7±14.1 |
| SpO2 at rest | 94.9±2.3 | 94.7±2.3 |
| Dyspnea at rest (mMRC) | 1.6±0.6 | 1.7±0.7 |
| 6-MWT distance | 406±118 | 375±101 |
| Desaturation after 6-MWR – number (%) | 90 (48) | 86 (46) |
| Dyspnea increase (Borg scale) after 6-MWT – number (%) | 104 (56) | 121 (64) |

\* Plus-minus values are means ± SD. FVC: forced vital capacity; SpO2: oxygen saturation; mMRC: modified medical research council dyspnea scale; 6-MWT: 6-minute walk test.

### Endpoints

In the FAS population, rate of predicted FVC recovery by Day 71 (primary endpoint) was 10.7% in the group of patients receiving BA and 10.4% in the placebo group (diff=0.32%; 95%CI: -2.22– 2.85; p=0.8). By Day 180, predicted FVC increased by 14.6% and 13.3% in the BA and placebo groups, respectively (dif=1.27%; 95%CI: -1.48–3.97; p=0.35). BA also showed a trend towards an increase in the proportion of patients achieving ≥10% ppFVC growth. On Day 180, 68% of patients in the BA group and 58.4% of patients in the placebo group exhibited at least a 10% ppFVC increase from baseline (OR=1.51; 95%CI: 0.99–2.32; p=0.058: Fig. 2).

**Figure 2.**
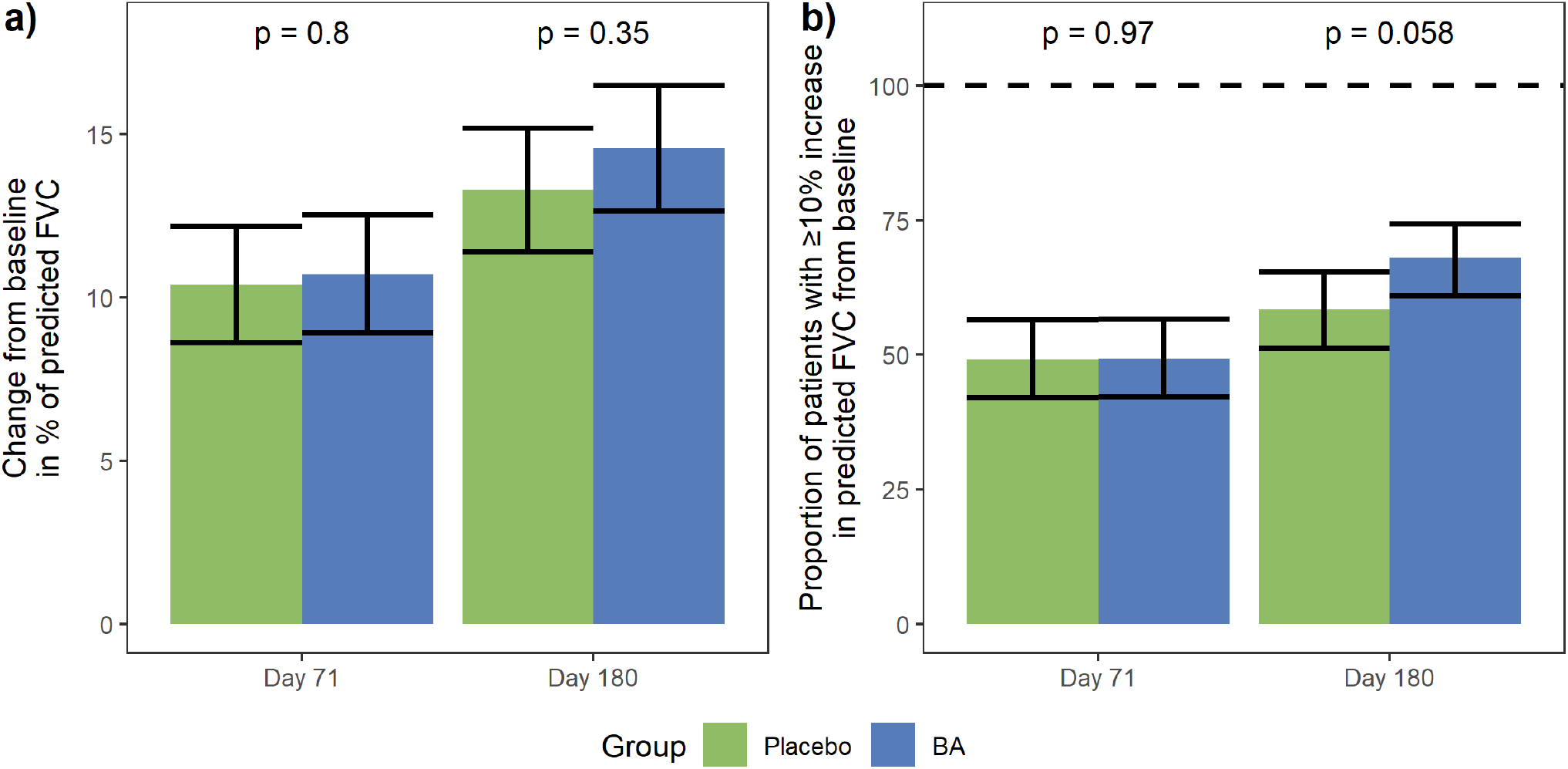
a) Change from baseline in percent of predicted forced vital capacity (FVC) by Day 71 (primary endpoint) and Day 180. b) Proportion of patients achieving at least 10% increase in predicted FVC by Day 71 and Day 180.

Review of the secondary endpoints data indicates that exposure to BA was associated with a marked increase in exercise tolerance. By Day 71, 4.7% of patients in the BA group versus 12.4% of patients in the placebo group exhibited desaturation after the 6-MWT, indicating a 62% reduction in exertional desaturation (OR=0.35; 95%CI: 0.16–0.71; p=0.0051). By Day 71, 28.8% of patients in the BA group and 39.4% of patients in the placebo group had exertional dyspnea after the 6-MWT (OR=0.62; 95%CI: 0.39–0.98; p=0.043). The difference in exertional dyspnea was maintained at Day 180: 23.7% of patients in the BA group versus 37.3% of patients in the placebo group exhibited dyspnea increase after the 6-MWT (OR=0.52; 95%CI: 0.33–0.83; p=0.0058). BA was linked to an apparent decrease in dyspnea at rest. On Day 180, dyspnea had decreased by ≥1 point on the mMRC scale in 82% of patients in the BA group and in 73.6% of the placebo group (OR=1.64; 95%CI: 1.00–2.70; p=0.051; Fig. 3).

**Figure 3.**
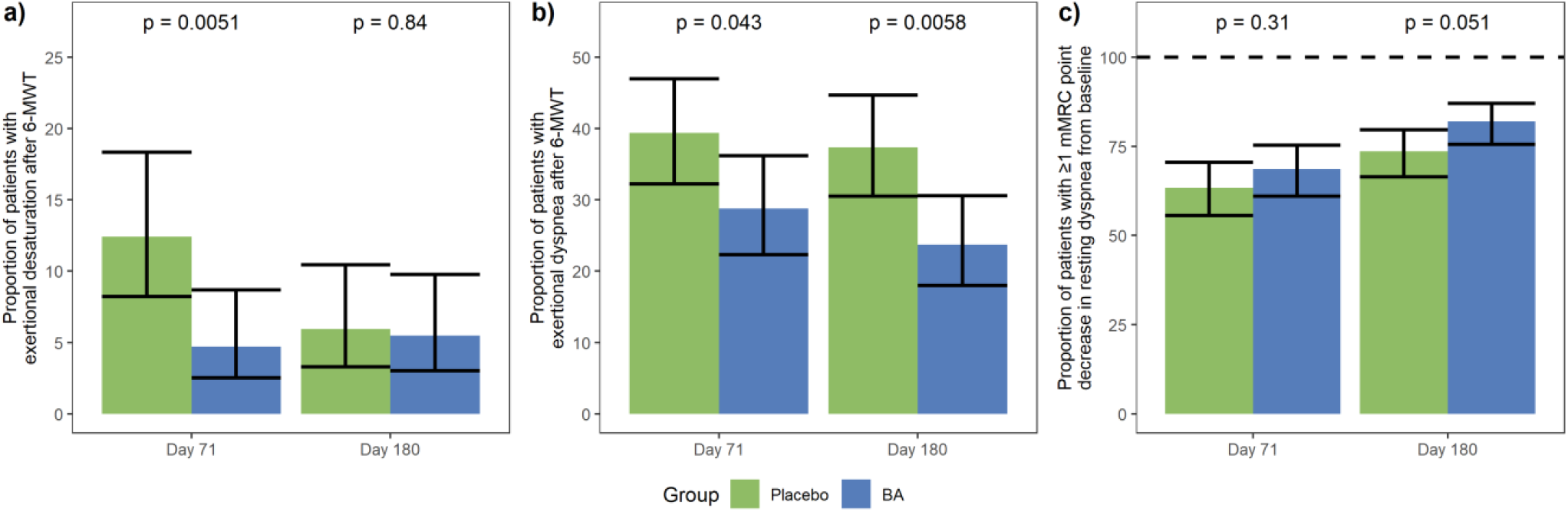
a) Proportion of patients with exertional desaturation defined as a ≥4% decrease in SpO2 readings after a six-minute walk test (6-MWT) on Day 71 and Day 180. b) Proportion of patients with exertional dyspnea defined by a ≥2 point increase on the Borg scale after 6-MWT on Day 71 and Day 180. c) Proportion of patients achieving a ≥1 point decrease in resting dyspnea on the modified Medical Research Council scale (mMRC) at Day 71 and Day 180.

There was no significant difference in resting SpO2, the proportion of patients achieving ≥50m increase in 6-MWT distance, cough severity, or quality of life assessment between the BA and the placebo groups (Supplementary appendix Table S1 and Fig. S1).

### Exploratory Analysis

Sub-population analysis demonstrated that BA was associated with a significantly higher FVC growth rate versus placebo in patients with earlier COVID-19 infection, and with cardiovascular comorbidities.

In patients who had COVID-19 before February 14, 2022 (median date), BA treatment was linked to a 15.9% ppFVC increase by Day 180, vs. 11.7% in the placebo group (n=188; diff=4.17%; 95%CI: 0.63–7.71; p=0.021). In patients with underlying cardiovascular conditions, ppFVC increased by 16% in the BA group by Day 180 vs. 12.7% increase in the placebo group (n=281; diff=3.31%; 95%CI: 0.11–6.50; p=0.042). Interestingly, in patients without cardiovascular comorbidities BA was associated with slower ppFVC growth (n=95; diff=-4.89%; 95%CI: -9.77–-0.003; p=0.05).

In patients >60 years, 75.3% of BA patients and 58.3% of placebo patients reached ≥10% increase in ppFVC by Day 180 (n=195; OR=2.18; 95%CI: 1.16–4.19; p=0.017). In female patients, the corresponding values for the BA and placebo groups were 79.9% and 63.4%, respectively (n=229; OR=2.30; 95%CI: 1.27–4.25; p=0.0069: Fig. 4).

**Figure 4.**
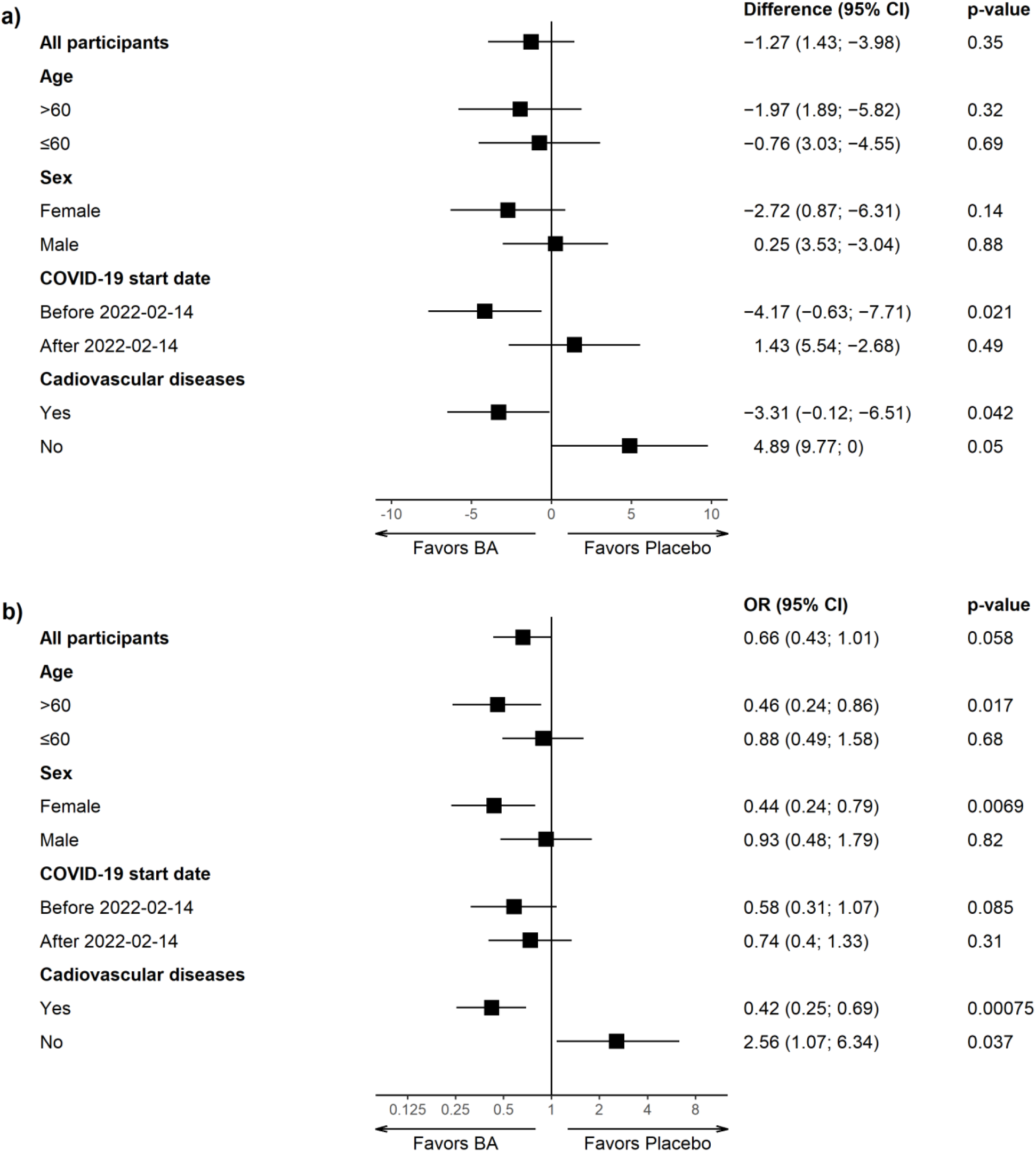
a) Forest plot of subgroup analysis of ppFVC change from the baseline by Day 180. b) Forrest plot of subgroup analysis of the proportion of patients achieving at least 10% increase in predicted FVC by Day 180.

In the FAS population BA demonstrated a trend towards an increase in FVC measured in milliliters rather than percentages of the predicted value. On Day 180, FVC increased by 519mL from the baseline in BA group and by 436mL in the placebo group (diff=83 mL; 95%CI: -13–179; p=0.093: Supplementary appendix Fig. S2).

Mean dyspnea at rest decreased by 0.95 points on the mMRC scale in the BA group and by 0.85 points in the placebo group at Day 180 (diff=-0.14; 95%CI: -0.27–-0.16; p=0.032). The average 6-MWT distance increased by 69.5m in the BA group and by 58.7m in the placebo group (diff=10.8m; 95%CI=-1.05 –22.67; p=0.072: Supplementary appendix Fig. S3).

### Safety

Seventy-three (37%) of 196 patients in the BA group and 49 (25%) of 196 patients in the placebo group experienced adverse events during the study. Eight adverse events in seven patients were deemed related to BA. These included three injection site reactions, and single cases of elevated alanine aminotransferase levels, elevated C-reactive protein, thrombocytopenia, hives, and angina pectoris. One participant receiving placebo experienced a case of thrombocytopenia which was considered related to treatment. Three patients in the BA group had serious adverse events, though these were not related to BA. Adverse events led to the discontinuation of the study drug in five patients receiving BA and one patient in the placebo group (Table 2).

**Table 2.** Adverse events.

| **Event** | **BA** | **Placebo** |
| --- | --- | --- |
|  | *number of patients (%)* |  |
| Any adverse events | 73 (37.2) | 49 (25) |
| Most frequent adverse events |  |  |
| - Respiratory tract infection | 11 (5.6) | 13 (6.6) |
| - COVID-19 | 4 (2) | - |
| Severe adverse event | 3 (1.5) | - |
| - Gastroenteritis | 1 (0.5) | - |
| - COVID-19 | 1 (0.5) | - |
| - Cholecystitis | 1 (0.5) | - |
| Adverse event leading to discontinuation of study treatment | 5 (2.5) | 1 (0.5) |
| - COVID-19 | 3 (1.5) | 1 (0.5) |
| - Candidiasis | 1 (0.5) | - |
| - Gastrointestinal diseases | 1 (0.5) | - |
| Adverse event considered related to study drug | 7 (3.6) | 1 (0.5) |
| - Injection reaction in 3 patients and hives in 1 patient | 3 (1.5) | - |
| - Elevated C-reactive protein | 1 (0.5) | - |
| - Elevated alanine aminotransferase | 1 (0.5) | - |
| - Thrombocytopenia | 1 (0.5) | 1 (0.5) |
| - Angina pectoris | 1 (0.5) | - |

## Discussion

We conducted a randomized, double-blind, placebo-controlled clinical trial to investigate the impact of BA on COVID-19 pulmonary sequelae in patients with a history of infection and residual lung abnormalities, restrictive pulmonary disease, dyspnea, and decreased oxygen saturation at rest or after exercise. The study did not achieve its primary endpoint of a significant change from the baseline in ppFVC by Day 71. However, we observed a significant BA effect on improving the exertional desaturation and dyspnea, and a marked FVC increase on Day 180 in specific subpopulations, 109 days after BA discontinuation. The rationale for conducting a clinical trial of BA stems from the potential role of HA in the pathogenesis of pulmonary sequelae of COVID-19. HA has been implicated in both fibrotic diseases and systemic inflammation, which are key features of COVID-19 pulmonary complications and long COVID. Several studies support targeting HA to treat COVID-19 and fibrotic diseases. Hyaluronidase-based interventions such as BA alleviated the development of bleomycin-induced lung fibrosis in mice [16]. Moreover, a recent open-label study on patients with pulmonary sequelae after COVID-19 revealed that treatment with BA was associated with marked improvements in pulmonary function, exercise tolerance, and respiratory symptoms.

On days 71 and 180, BA had no statistically significant effect on the ppFVC recovery rate, although by day 180, ppFVC increased slightly more in the BA group than in the placebo group (14.6% vs. 13.3%, p=0.35). Interestingly, this effect becomes more pronounced if FVC is measured in milliliters instead of as a percentage of predicted values, with FVC increasing from the baseline by an additional 83 ml in the BA group compared to the placebo group by Day 180 (p=0.093). By Day 180, BA also demonstrated a strong trend towards increasing the percentage of patients with a 10% ppFVC increase (p=0.058).

The possible trend towards FVC increase at Day 180, coupled with significant effects on FVC observed in several sub-populations by this time point, suggests that a longer treatment duration and later endpoint date might yield different results. However, the rationale behind the selected treatment duration and endpoint measurement date was intended to balance drug exposure with patient compliance and recovery rate. The 71-day treatment period was based on approved duration of BA use and on compliance considerations, as patients needed to visit the study site every five days for injections. Selection of Day 71 for endpoint measurement was based on existing knowledge about the resolution rate of restrictive lung disease post-COVID-19 [17]. This approach may not have fully captured the benefits of BA, as by Day 180, the mean FVC was 82% of the predicted value, indicating room for further growth.

The study included patients within an 11-month window between COVID-19 diagnosis and randomization, resulting in participants contracting SARS-CoV-2 over a 9-month period. According to epidemiological data, during this period, patients were likely infected with Delta, Omicron B1, B2, and B5 variants [18]. Enrollment was also agnostic to preceding COVID-19 severity and hospitalization status, leading to high population heterogeneity that might have contributed to the study failing to achieve its primary endpoint. Indeed, the study demonstrated a significant BA effect on FVC in patients who had earlier COVID-19.

Bovhyaluronidase azoximer was associated with an increase in exercise tolerance, seeing the proportion of patients exhibiting desaturation and dyspnea after exercise decrease by 62% and 27%, respectively. Exercise intolerance, not limited to patients with pulmonary sequelae, is one of the most common and persistent symptoms of long COVID-19 [2]. The effect we observed might be attributed to both pulmonary and extrapulmonary mechanisms. BA may have enhanced gas exchange by alleviating fibrosis and inflammation in the lungs. Alternatively, BA may have exerted anti-inflammatory effects at a systemic level, positively affecting cardiac function and vascular pathology [19, 20]. The absence of BA’s effect on pulmonary function in the FAS population further supports the extrapulmonary origin of increase in exercise tolerance. These findings underscore the need for further research on the impact of BA on exercise tolerance in long COVID patients, regardless of pulmonary function impairment. Interestingly, exercise intolerance is also a hallmark symptom of myalgic encephalomyelitis/chronic fatigue syndrome (ME/CFS), which has mechanistic similarities to long COVID, suggesting the potential of HA-targeting therapies for this condition [21, 22].

Patients with underlying cardiovascular diseases (CVDs) receiving BA demonstrated significantly shorter recovery of pulmonary function compared to patients receiving a placebo, as indicated by the change in FVC from baseline by day 180. Conversely, in patients without cardiovascular comorbidities, BA use was associated with a slower pace of FVC growth. These findings may be attributed to endothelial dysfunction and chronic inflammation, which are common mechanisms in CVDs and post-COVID-19 pulmonary sequelae [23,24,25].

Endothelial dysfunction starts with the shedding of the HA-containing endothelial glycocalyx, followed by an increase in both the synthesis of high-molecular-weight HA (HMW-HA) aimed at restoring the damaged glycocalyx layer and its degradation into pro-inflammatory low-molecular-weight HA (LMW-HA) [26]. Given that CVDs are associated with endothelial dysfunction, we suggest that in this study, the HA equilibrium was shifted towards LMW-HA in patients with cardiovascular comorbidities compared to those without such conditions. This shift may explain the opposing effects of BA on these two subgroups.

In this study, patients with cardiovascular comorbidities likely exhibited greater levels of inflammation compared to those without cardiovascular diseases [24]. We therefore suggest that the accelerated recovery of pulmonary function specific to patients with cardiovascular diseases can be explained by the established anti-inflammatory effects of hyaluronidases and HA-synthase inhibitors [27]. Studies demonstrating the beneficial effect of hyaluronidases in cardiovascular pathology further support this explanation [28].

BA significantly increased FVC recovery rate by Day 180 in patients who had COVID-19 earlier than the median date for preceding infection and did not affect pulmonary function in those infected later. Epidemiological data indicate that two-thirds of patients in the first subgroup were infected with the Delta variant of SARS-CoV-2, whereas the remaining patients encountered the Omicron BA1 and BA1 variants [18]. Patients in the second subgroup were likely infected with the Omicron B1, B2, and B5 variants. In comparison to the Omicron variants, the Delta variant is linked to more severe COVID-19, which is a risk factor for developing pulmonary sequelae [29,30].

In female patients and those aged >60 years, the use of BA significantly increased the proportion of participants achieving a ≥10% growth in ppFVC. Both female sex and advanced age are recognized as risk factors for long COVID-19, with female sex also being associated with the persistence of symptoms [31, 32]. We suggest that patients infected with earlier versions of the virus, female patients, and those aged over 60 years developed a more severe long COVID phenotype that was responsive to BA treatment.

The study has several limitations. The choice of 71-day treatment duration and Day 71 for primary outcome measurement was likely not optimal for assessing the effect of BA on FVC recovery in studied population, as discussed above. The absence of DLCO testing limits our understanding of pulmonary function dynamics. The study did not measure fatigue, a clinically relevant long COVID symptom linked to exercise intolerance. In addition, the study did not test for markers of inflammation and vascular damage which could have provided mechanistic explanations for the observed effects of BA.

In conclusion, we found that BA significantly increased exercise tolerance in patients with long-term pulmonary sequelae of COVID-19. BA also significantly accelerated pulmonary function recovery in patients with cardiovascular comorbidities, patients infected with earlier variants of SARS-CoV-2, female patients and patients aged > 60 years. However, it did not affect pulmonary function recovery in the overall population. The differential effects of BA on patients with and without cardiovascular comorbidities highlight the antagonistic roles of HA in post-COVID-19 pulmonary sequelae and suggest that HA-targeting therapies require careful selection of study populations. The study findings also suggest that longer exposure to hyaluronidase may be necessary to impact on lung function recovery. Overall, our findings support the further investigation of hyaluronidase-based therapies for long COVID, with or without pulmonary sequelae.

## Data Availability

The data forms a part of an ongoing research programme. Authors have the intention is to make the data publicly available once the programme is complete.

