## Supplementary Material for "Bovhyaluronidase azoximer for long-term pulmonary sequelae of COVID-19: a randomized, double-blind, placebo-controlled trial"

**Table S1. Primary and secondary endpoints**

| **Continuous Endpoints*** | **BA** | **Placebo** | **Diff (CI 95%)** | ***p*-value** |
| --- | --- | --- | --- | --- |
| Change from baseline in percent predicted FVC | |  |  |  |
| - Day 71 *primary endpoint* | 10.71 (8.91 to 12.52) | 10.40 (8.61 to 12.18) | 0.32 (-2.22 to 2.85) | 0.80 |
| - Day 180 | 14.57 (12.65 to 16.49) | 13.30 (11.40 to 15.20) | 1.27 (-1.43 to 3.98) | 0.35 |
| Change from baseline in resting SpO2 | |  |  |  |
| - Day 71 | 1.90 (1.67 to 2.12) | 1.71 (1.49 to 1.93) | 0.19 (-0.13 to 0.50) | 0.24 |
| - Day 180 | 2.11 (1.89 to 2.34) | 2.03 (1.81 to 2.25) | 0.08 (-0.23 to 0.39) | 0.60 |
| Change from baseline in cough severity (VAS) | |  |  |  |
| - Day 71 | -17.04 (-20.24 to -13.84) | -14.12 (-17.33 to -10.90) | -2.92 (-7.46 to 1.62) | 0.21 |
| - Day 180 | -19.56 (-22.71 to -16.41) | -15.91 (-19.08 to -12.73) | -3.65 (-8.13 to 0.82) | 0.11 |
| Change from baseline in quality of life assessed by European Quality of Life 5-Dimension 5-Level Questionnaire | | | | |
| - Day 71 | 11.56 (9.88 to 13.24) | 12.05 (10.39 to 13.71) | -0.49 (-2.86 to 1.88) | 0.69 |
| - Day 180 | 16.19 (14.47 to 17.92) | 15.02 (13.32 to 16.73) | 1.17 (-1.26 to 3.61) | 0.35 |
| **Binary endpoints**† | **BA** | **Placebo** | **OR (CI 95%)** | ***p*-value** |
| Proportion of patients achieving at least 10% increase in predicted FVC | | |  |  |
| - Day 71 | 49.36 (42.11 to 56.63) | 49.19 (42.02 to 56.40) | 1.01 (0.67 to 1.52) | 0.97 |
| - Day 180 | 68.02 (60.89 to 74.40) | 58.45 (51.20 to 65.35) | 1.51 (0.99 to 2.32) | 0.058 |
| Proportion of patients with SpO2 at rest ≥ 93% | | |  |  |
| - Day 71 | 99.65 (97.09 to 99.96) | 99.05 (95.98 to 99.78) | 2.73 (0.34 to 56.00) | 0.39 |
| - Day 180 | 99.47 (96.29 to 99.93) | 98.95 (95.89 to 99.74) | 1.99 (0.19 to 43.15) | 0.58 |
| Proportion of patients with exertional desaturation after 6-MWT‡ | | |  |  |
| - Day 71 | 4.74 (2.51 to 8.66) | 12.41(8.21 to 8.66) | 0.35 (0.16 to 0.71) | 0.0051 |
| - Day 180 | 5.49 (3.02 to 9.77) | 5.93 (3.30 to 10.44) | 0.92 (0.41 to 0.71) | 0.84 |
| Proportion of patients achieving ≥50 m increase from baseline in distance walked in 6-MWT | | | |  |
| - Day 71 | 38.98 (32.05 to 46.39) | 33.92 (27.39 to 41.14) | 1.24 (0.81 to 1.92) | 0.32 |
| - Day 180 | 56.27 (48.78 to 63.47) | 48.42 (41.14 to 55.76) | 1.37 (0.90 to 2.10) | 0.14 |
| Proportion of patients with exertional dyspnea after 6-MWT§ | | | |  |
| - Day 71 | 28.76 (22.32 to 36.20) | 39.37 (32.22 to 47.01) | 0.62 (0.39 to 0.98) | 0.043 |
| - Day 180 | 23.73 (18.01 to 30.6) | 37.33 (30.52 to 44.68) | 0.52 (0.33 to 0.83) | 0.0058 |
| Proportion of patients achieving a ≥1 mMRC point decrease in resting dyspnea | | |  |  |
| - Day 71 | 68.64 (61.06 to 75.34) | 63.38 (55.65 to 70.48) | 1.26 (0.80 to 2.00) | 0.31 |
| - Day 180 | 82.00 (75.60 to 87.00) | 73.57 (66.45 to 79.64) | 1.64 (1.00 to 2.70) | 0.051 |
| * Changes from baseline are ANCOVA means (CI 95%);†Proportions are GLM means (CI 95%); ‡Defined as a ≥4% decrease in SpO2 readings after a six-minute walk test (6-MWT); §Defined by a ≥2 point increase on the Borg scale | | | | |

**Figure S1.** a) Change from baseline in resting SpO2 by Day 71 and Day 180. b) Change from the baseline in cough severity (VAS) by Day 71 and Day 180. c) Proportion of patients with resting SpO2>93%. d) Proportion of patients achieving ≥50 m increase from baseline in distance walked in six-minute walk test (6-MWT). e) Proportion of patients achieving a ≥1 point decrease in resting dyspnea on the modified Medical Research Council scale (mMRC) at Day 71 and Day 180. f) Change from the baseline in quality of life assessed by European Quality of Life 5-Dimension 5-Level Questionnaire (Euro-QoL-5D-5L).

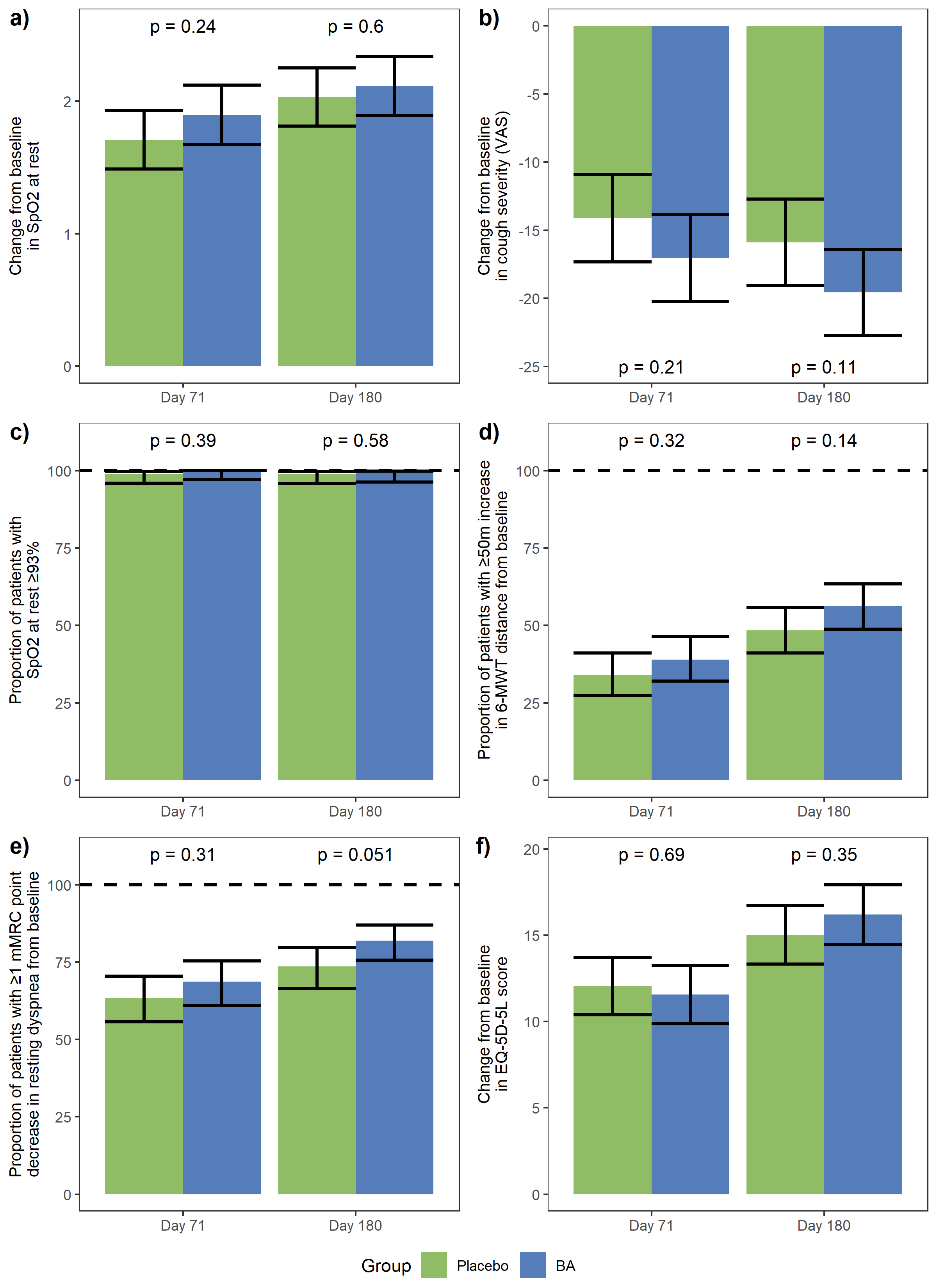

**Figure S2.** a) Change from baseline in dyspnea at rest assessed by the modified Medical Research council scale (mMRC) by Day 71 and Day 180. b) Change from the baseline in distance walked during the six-minute walk test (6-MWT) by Day 71 and Day 180.

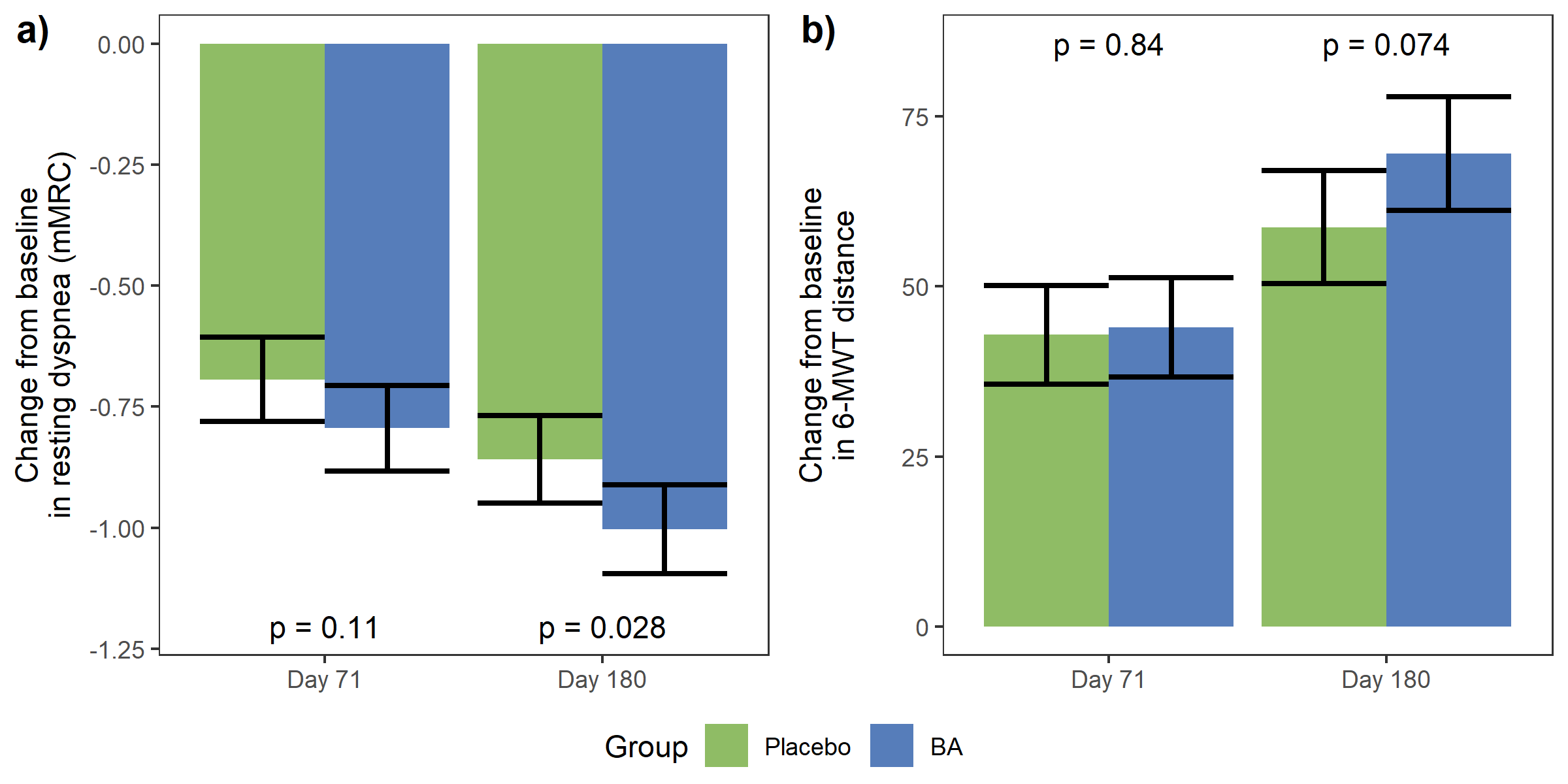

**Figure S3** a) Change from baseline in forced vital capacity (FVC) measured in milliliters by Day 71 Day 180. b) Proportion of patients achieving at least 10% increase in FVC (ml) by Day 71 and Day 180.

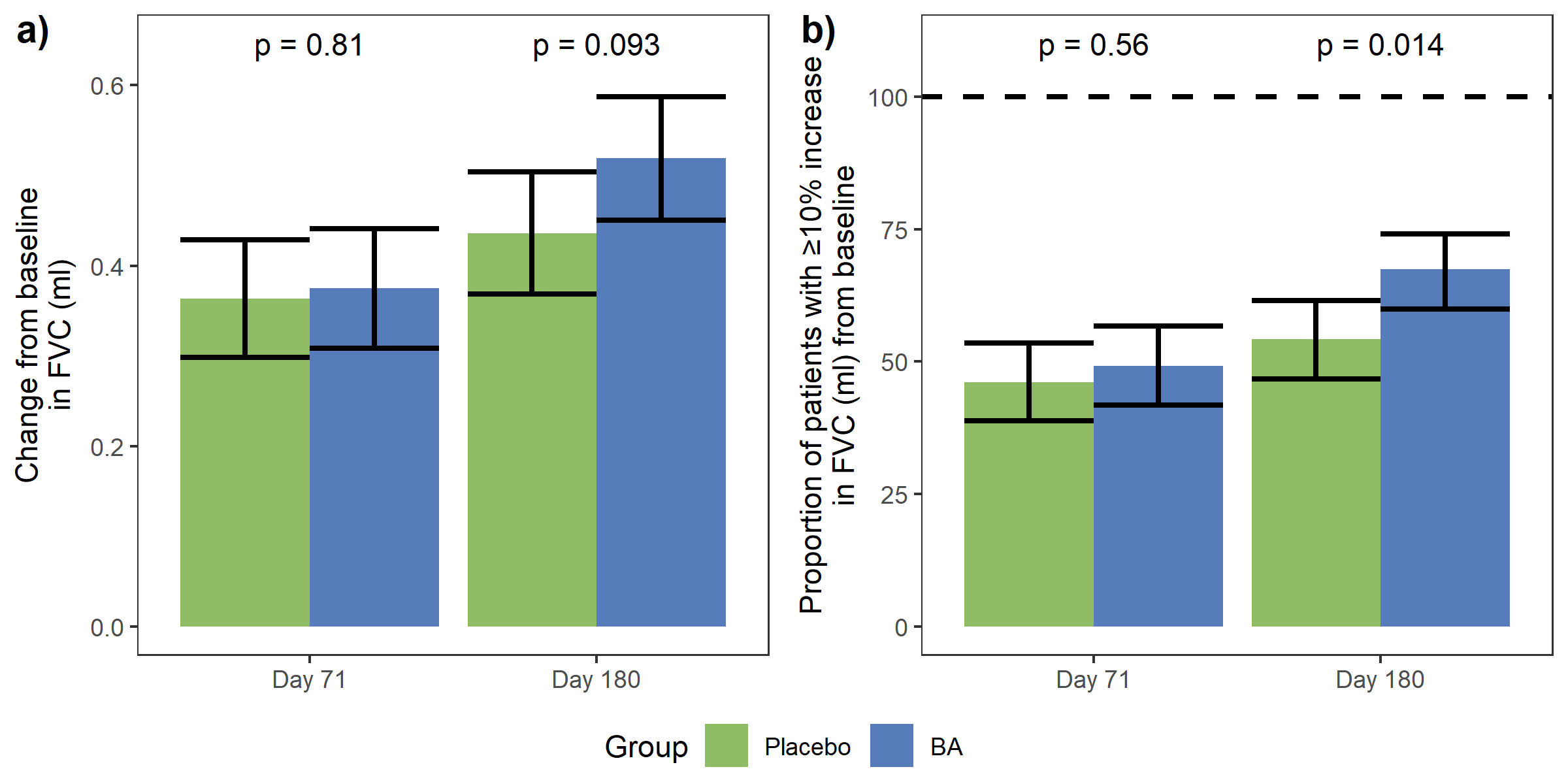

**List of study sites**

1. Regional Clinical Hospital No. 3, State Budgetary Healthcare Institution, Chelyabinsk, Russia
2. Kuzbass Clinical Emergency Medical Hospital named after M.A. Podgorbunsky, State Autonomous Healthcare Institution, Kemerovo, Russia
3. Medical Center "Reavita Med SPb"; Therapy Department, LLC, St. Petersburg, Russia
4. Medical Center for Diagnostics and Prevention Plus, LLC, Yaroslavl, Russia
5. Consultative and Diagnostic Center No. 85, St. Petersburg State Budgetary Healthcare Institution, St. Petersburg, Russia
6. Siberian State Medical University, Federal State Budgetary Educational Institution of Higher Education, Ministry of Health of the Russian Federation, Tomsk, Russia
7. Northern Medical Clinical Center named after N.A. Semashko, Federal State Budgetary Healthcare Institution, Federal Medical-Biological Agency, Arkhangelsk, Russia
8. Kirov State Medical University, Federal State Budgetary Educational Institution of Higher Education, Ministry of Health of the Russian Federation, Kirov, Russia
9. First Moscow State Medical University named after I. M. Sechenov, Federal State Autonomous Educational Institution of Higher Education, Ministry of Health of the Russian Federation; University Clinical Hospital No. 4, Moscow, Russia
10. Alliance Biomedical-Ural Group, LLC, Izhevsk, Russia
11. National Medical Research Center for Phthisiopulmonology and Infectious Diseases, Federal State Budgetary Healthcare Institution, Ministry of Health of the Russian Federation; Ural Research Institute of Phthisiopulmonology, Yekaterinburg, Russia
12. Kazan State Medical University, Federal State Budgetary Educational Institution of Higher Education, Ministry of Health of the Russian Federation, Kazan, Russia
13. UNIMED-S, CJSC, Moscow, Russia
14. Voronezh Regional Clinical Hospital No. 1, Budgetary Healthcare Institution of the Voronezh Region, Voronezh, Russia
15. Research Center Eco-Safety, LLC, St. Petersburg, Russia
16. Clinical Hospital "RZD-Medicine" of the City of Chelyabinsk, Private Healthcare Institution, Chelyabinsk, Russia
17. Medical Center "Reavita Med SPb"; Rehabilitation Department, LLC, St. Petersburg, Russia
18. Ivanovo State Medical Academy, Federal State Budgetary Educational Institution of Higher Education, Ministry of Health of the Russian Federation, Kakhma, Russia
19. National Medical Research Center for Therapy and Preventive Medicine, Federal State Budgetary Healthcare Institution, Ministry of Health of the Russian Federation, Moscow, Russia
20. Clinical Hospital No. 9, State Autonomous Healthcare Institution of the Yaroslavl Region, Yaroslavl, Russia
21. Energy of Health, LLC, St. Petersburg, Russia
22. Engels City Clinical Hospital No. 1, State Autonomous Healthcare Institution, Engels, Russia
23. Astarta, LLC, St. Petersburg, Russia
24. City Consultative and Diagnostic Center No. 1, St. Petersburg State Budgetary Healthcare Institution, St. Petersburg, Russia
25. Regional Clinical Hospital, State Healthcare Institution, Saratov, Russia
26. Professor's Clinic, LLC, Perm, Russia
27. RC Medical, LLC, Novosibirsk, Russia
28. Consultative and Diagnostic Center with Polyclinic, Federal State Budgetary Institution, Presidential Administration of the Russian Federation, St. Petersburg, Russia
29. Central Research Institute of Epidemiology, Federal Budgetary Institution of Science, Federal Service for Surveillance on Consumer Rights Protection and Human Wellbeing, Moscow, Russia
30. X7 Clinical Research, LLC, St. Petersburg, Russia
31. Group of Companies Persona, LLC, Nizhny Novgorod, Russia
32. Aramil City Hospital, State Autonomous Healthcare Institution of the Sverdlovsk Region, Aramil, Russia
33. Russian Scientific Center of Surgery named after Academician B.V. Petrovsky, Federal State Budgetary Scientific Institution, Moscow, Russia
34. Clinical Hospital No. 1, Regional State Budgetary Healthcare Institution, Smolensk, Russia
35. Saratov State Medical University named after V.I. Razumovsky, Federal State Budgetary Educational Institution of Higher Education, Ministry of Health of the Russian Federation; University Clinical Hospital No. 1 named after S.R. Mirotvortsev, Saratov, Russia
36. Hospital for War Veterans of Kazan, State Autonomous Healthcare Institution, Kazan, Russia
37. Medical Center "Revma-Med", LLC, Kemerovo, Russia

**Exploratory analysis methods**

Patients were stratified by characteristics of their previous COVID-19, by demographic characteristics, and by the severity of the patients' conditions at baseline. Specifically, patients were stratified by COVID-19 severity (mild and moderate vs. severe and very severe) and hospitalization status; by COVID-19 start date (before February 14, 2022, vs. on or after February 14, 2022) and time from COVID-19 start date to randomization (≤ 223 days vs. > 223 days); by age (≤ 60 years vs. > 60 years), sex, and BMI (≤ 28.8 vs. > 28.8 kg/m2); by the presence of cardiovascular comorbidities defined by MedDRA system organ classes; by ppFVC (≤ 70.6% vs. > 70.6%) and percentage of lung damage (≤ 15% vs. > 15%) at screening; by baseline SpO2 at rest (≤ 94% vs. > 94%); and by baseline distance in the 6-MWT (≤ 382 meters vs. > 382 meters). Stratification boundaries of continuous variables were set at median values to ensure similar sizes of subpopulations, with the exception of baseline SpO2 and percentage of lung damage.

Following outcomes not specified in the study protocol were included in the explanatory analysis: change from baseline in FVC (ml) by days 71 and 180; change from baseline in resting dyspnea (mMRC) by days 71 and 180; and change from baseline in 6-MWT distance by days 71 and 180.

**Sample size calculation**

The sample size for the primary endpoint analysis was calculated to be 352 patients, with 176 patients per treatment group, based on a Cohen's d effect size of 0.3, 80% power, and a two-sided significance level of 5%. Cohen’s coefficient was estimated using the BA effect on the change in ppFVC from baseline in the pilot, open-label study (17). To account for an estimated 15% screening failure rate and a 10% dropout rate after randomization, it was decided to randomize 392 patients (196 per group) and to screen up to 461 patients.

**Randomization, blinding and allocation concealment**

Eligible patients were randomized to receive either BA 3000 U once every 5 days or a matching placebo in a 1:1 allocation ratio, generated by the Electronic Data Capture/Integrated Web-Based Randomization System (EDC/IWRS). BA and the placebo had identical packaging with unique identifiers corresponding to the treatment group. The list of identifiers was available only to the unblinded specialist at the clinical research organization. Study investigators and patients were blinded. An unblinded staff member from the investigator team at each study site was responsible for the registration, storage, and dispensation of study treatment packages. The unblinded staff member was not involved in the filing of source documents, completion of case report forms, or observation of patients.

**Study funding**

Study was funded by NPO Petrovax Pharm LLC. BA and matching placebo were produced and supplied by NPO Petrovax Pharm LLC.

**Ethics declaration**

The study was independently approved by the following ethics committees:

1. Ethics Committee of the Russian Ministry of Health, Moscow, Russia
2. Local Independent Ethics Committee of Regional Clinical Hospital No. 3, State Budgetary Healthcare Institution, Chelyabinsk, Russia
3. Independent Ethics Committee of Kuzbass Clinical Emergency Medical Hospital named after M.A. Podgorbunsky, State Autonomous Healthcare Institution, Kemerovo, Russia
4. Local Ethics Committee of the Medical Center "Reavita Med SPb," Therapy Department, LLC, St. Petersburg, Russia
5. Ethics Committee of the Medical Center for Diagnostics and Prevention Plus, LLC, Yaroslavl, Russia
6. Ethics Committee of the Consultative and Diagnostic Center No. 85, St. Petersburg State Budgetary Healthcare Institution, St. Petersburg, Russia
7. Independent Ethics Committee of the Siberian State Medical University, Federal State Budgetary Educational Institution of Higher Education, Ministry of Health of the Russian Federation, Tomsk, Russia
8. Ethics Committee of the Northern Medical Clinical Center named after N.A. Semashko, Federal State Budgetary Healthcare Institution, Federal Medical-Biological Agency, Arkhangelsk, Russia
9. Local Ethics Committee of Kirov State Medical University, Federal State Budgetary Educational Institution of Higher Education, Ministry of Health of the Russian Federation, Kirov, Russia
10. Local Ethics Committee of the First Moscow State Medical University named after I. M. Sechenov, Federal State Autonomous Educational Institution of Higher Education, Ministry of Health of the Russian Federation; University Clinical Hospital No. 4, Moscow, Russia
11. Local Ethics Committee of Alliance Biomedical-Ural Group, LLC, Izhevsk, Russia
12. Ethics Committee of the National Medical Research Center for Phthisiopulmonology and Infectious Diseases, Federal State Budgetary Healthcare Institution, Ministry of Health of the Russian Federation; Ural Research Institute of Phthisiopulmonology, Yekaterinburg, Russia
13. Local Ethics Committee of Kazan State Medical University, Federal State Budgetary Educational Institution of Higher Education, Ministry of Health of the Russian Federation, Kazan, Russia
14. Local Ethics Committee of UNIMED-S, CJSC, Moscow, Russia
15. Ethics Committee of Voronezh Regional Clinical Hospital No. 1, Budgetary Healthcare Institution of the Voronezh Region, Voronezh, Russia
16. Ethics Committee of Research Center Eco-Safety, LLC, St. Petersburg, Russia
17. Ethics Committee of the Clinical Hospital "RZD-Medicine" in Chelyabinsk, Private Healthcare Institution, Chelyabinsk, Russia
18. Local Ethics Committee of Medical Center "Reavita Med SPb," Rehabilitation Department, LLC, St. Petersburg, Russia
19. Local Ethics Committee of Ivanovo State Medical Academy, Federal State Budgetary Educational Institution of Higher Education, Ministry of Health of the Russian Federation, Kakhma, Russia
20. Ethics Committee of the National Medical Research Center for Therapy and Preventive Medicine, Federal State Budgetary Healthcare Institution, Ministry of Health of the Russian Federation, Moscow, Russia
21. Local Ethics Committee of Clinical Hospital No. 9, State Autonomous Healthcare Institution of the Yaroslavl Region, Yaroslavl, Russia
22. Local Ethics Committee of Energy of Health, LLC, St. Petersburg, Russia
23. Ethics Committee of Engels City Clinical Hospital No. 1, State Autonomous Healthcare Institution, Engels, Russia
24. Local Ethics Committee of Astarta, LLC, St. Petersburg, Russia
25. Ethics Committee of the City Consultative and Diagnostic Center No. 1, St. Petersburg State Budgetary Healthcare Institution, St. Petersburg, Russia
26. Independent Ethics Committee of Regional Clinical Hospital, State Healthcare Institution, Saratov, Russia
27. Ethics Committee of Professor's Clinic, LLC, Perm, Russia
28. Ethics Committee of RC Medical, LLC, Novosibirsk, Russia
29. Ethics Committee of the Consultative and Diagnostic Center with Polyclinic, Federal State Budgetary Institution, Presidential Administration of the Russian Federation, St. Petersburg, Russia
30. Ethics Committee of the Central Research Institute of Epidemiology, Federal Budgetary Institution of Science, Federal Service for Surveillance on Consumer Rights Protection and Human Wellbeing, Moscow, Russia
31. Ethics Committee of X7 Clinical Research, LLC, St. Petersburg, Russia
32. Ethics Committee of the Group of Companies Persona, LLC, Nizhny Novgorod, Russia
33. Ethics Committee of Aramil City Hospital, State Autonomous Healthcare Institution of the Sverdlovsk Region, Aramil, Russia
34. Ethics Committee of the Russian Scientific Center of Surgery named after Academician B.V. Petrovsky, Federal State Budgetary Scientific Institution, Moscow, Russia
35. Ethics Committee of Clinical Hospital No. 1, Regional State Budgetary Healthcare Institution, Smolensk, Russia
36. Local Ethics Committee of Saratov State Medical University named after V.I. Razumovsky, Federal State Budgetary Educational Institution of Higher Education, Ministry of Health of the Russian Federation; University Clinical Hospital No. 1 named after S.R. Mirotvortsev, Saratov, Russia
37. Ethics Committee of the Hospital for War Veterans of Kazan, State Autonomous Healthcare Institution, Kazan, Russia
38. Ethics Committee of Medical Center "Revma-Med," LLC, Kemerovo, Russia
